# A Novel Dual-Guide CRISPR-Cas13 Strategy Improves Specificity for Single-Nucleotide Variant Detection

**DOI:** 10.1101/2025.09.24.25336526

**Authors:** Araceli Aguilar-González, Ismael Martos-Jamai, Iris Ramos-Hernández, Francisco Javier Molina-Estévez, Pilar Puig-Serra, Sandra Rodríguez-Perales, Raúl Torres, Rosario María Sánchez-Martín, Juan José Díaz-Mochón, Francisco Martín

## Abstract

The emergence of CRISPR-Cas systems has transformed nucleic acid detection and manipulation. Cas13, a type VI CRISPR effector, targets RNA with high sensitivity through both *cis* (target RNA) and *trans* (collateral RNA) cleavage. This property enables the use of fluorescent reporters for sensitive diagnostics. However, Cas13’s heightened sensitivity also leads to reduced specificity due to its susceptibility to single-nucleotide mismatches, potentially causing off-target effects. To overcome this limitation, we developed the first dual-guide RNA system for Cas13 that enhances mismatch discrimination and improves target specificity. This system employs two distinct RNAs—dcrRNA and dtracrRNA—which hybridise to refine target recognition and activation. *In vitro* experiments demonstrated robust cis- and trans-RNase activity, indicating efficient and specific cleavage. The system accurately detected SARS-CoV-2 RNA, demonstrating its potential for pathogen diagnostics, and successfully discriminated between KRAS G12D and G12C mutations—clinically relevant single-nucleotide variants in cancer diagnosis. These results highlight the dual-guide Cas13 platform’s potential for precise, rapid, and reliable RNA detection. Overall, this approach represents a significant advance over conventional Cas13 systems, offering improved specificity without compromising sensitivity. Its versatility makes it a promising tool for next-generation molecular diagnostics and precision gene editing applications.

**GRAPHICAL ABSTRACT:** 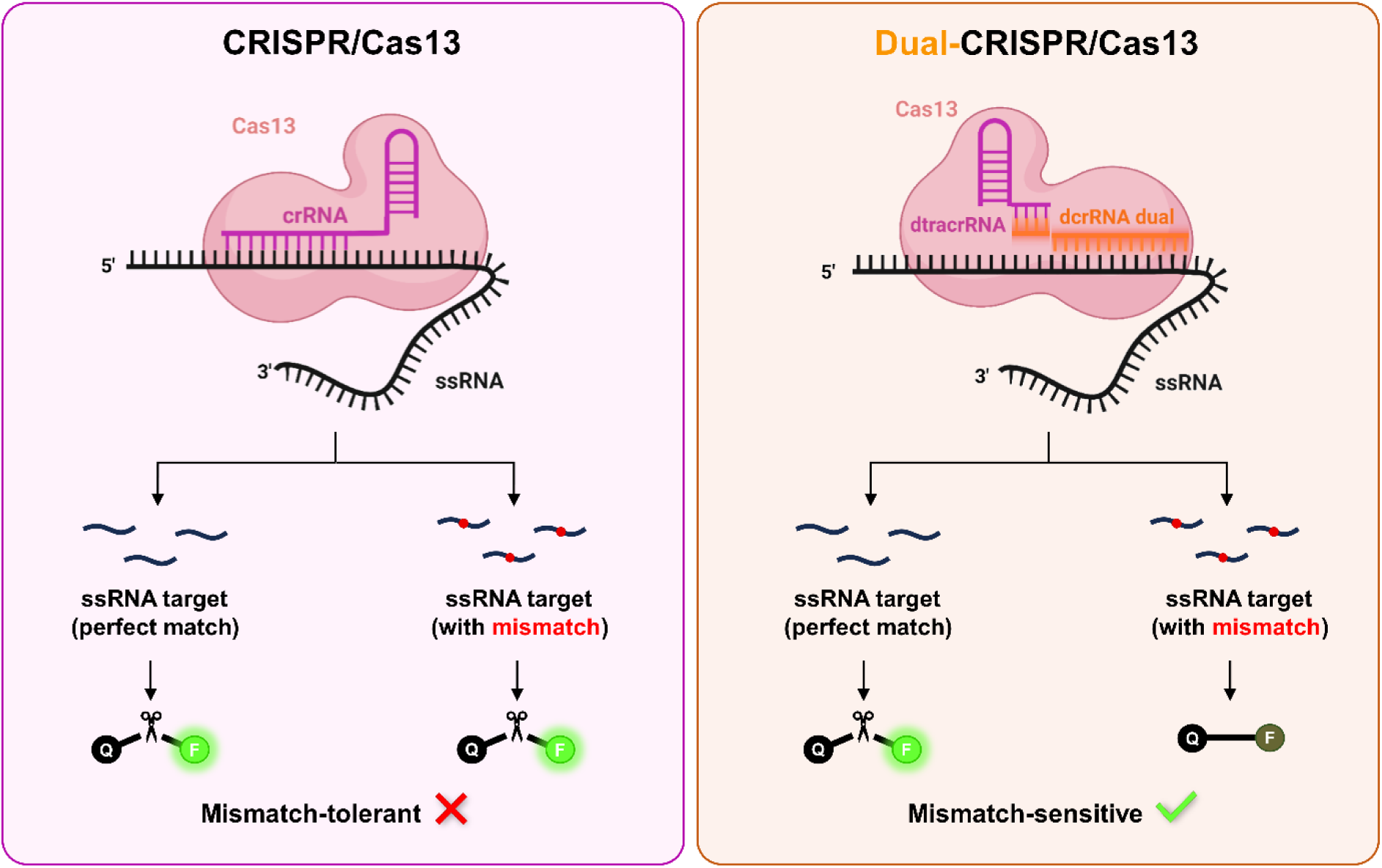

## INTRODUCTION

Molecular diagnostics based on nucleic acid detection have become a central component of modern medicine, enabling the identification of pathogens, genetic alterations, and transcriptional signatures across a range of biomedical applications [1, 2]. In particular, the ability to distinguish single-nucleotide variants (SNVs) is critical for differentiating closely related viral strains, detecting drug resistance mutations, or identifying oncogenic drivers (RAS, BRAF, EGFR, and other), as well as microRNAs [3–6]. Although PCR and hybridization-based techniques such as TaqMan probes, melting curve analysis and microarrays are widely used for SNV detection, they often suffer from limited mismatch discrimination, cross-reactivity, and inflexible design [7–9]. Sequencing-based approaches including next-generation sequencing (NGS) provide higher accuracy, especially for short RNA species, however its complexity and cost limit widespread implementation [9, 10]. For these reasons, alternative technologies that effectively balance sensitivity, specificity, and applicability are required.

Clustered Regularly Interspaced Short Palindromic Repeats (CRISPR) and CRISPR-associated (Cas) proteins have gained significant attention in recent years as promising alternatives. CRISPR/Cas systems have transformed the field of molecular biology, enabling precise and programmable nucleic acid targeting across a wide range of organisms and applications, including precision medicine, diagnostics, and biotechnology [11–13]. While early studies focused primarily on DNA-editing systems such as Cas9, increasing attention has been given to RNA-targeting effectors, particularly the class 2 type VI CRISPR effector Cas13. This enzyme is guided by a single RNA molecule to recognize and cleave the complementary target RNA (*cis* activity), while also inducing collateral (*trans*) RNase activity that results in non-specific degradation of surrounding RNAs [14, 15]. These properties have made Cas13 an attractive tool for transcriptome engineering, transient mRNA knockdown, and molecular diagnostics, where it has demonstrated high sensitivity in the nucleic acid-based detection of pathogenic organisms [16–19]. However, this sensitivity likewise results in a notable tolerance for single-nucleotide mismatches, raising concerns about off-target effects in both research and clinical settings. Consequently, enhancing mismatch discrimination in CRISPR/Cas13 platforms remains a critical challenge for their reliable use in high-specificity contexts [20–22].

To overcome these limitations, several strategies have been proposed to improve Cas13 mismatch discrimination and specificity. One common approach involves deliberately introducing synthetic base-pair alterations into the guide RNA in addition to the naturally occurring target mismatch, thus creating multiple mismatches (typically three or more) between guide and target sequences [23, 24]. This cumulative effect reduces off-target binding, leading to improved specificity. However, this strategy is highly empirical and target-specific, requiring careful optimization of the number, position, and nature of these mismatches, which restricts its applicability for routine use. Another strategy to improve off-target discrimination in Cas13 systems involves engineering the protein itself. This includes modifications to key residues in the catalytic domains or regions involved in crRNA binding, aiming to improve target recognition and reduce collateral activity. While these approaches can indeed improve specificity, they typically require extensive structural analysis, are labour-intensive, and often result in context-dependent performance [25, 26].

Recent studies have explored the viability of splitting the guide RNA in CRISPR systems beyond Cas9 [27–29]. Among the included studies, split CRISPR RNA configurations for Cas12a were able to reconstitute functional complexes *in vitro* under optimized conditions. However, these approaches have been limited to the detection of short RNA targets (such as microRNAs) due to the constrained guide length (typically less than 20 nucleotides). Achieving detectable activity often requires elevated concentrations of both protein and guide RNAs [29]. Because Cas12a is inherently a DNA-targeting enzyme, its direct application to RNA detection remains limited [30]. This restricts the use of Cas12a-based split systems in diagnostic settings involving longer or more complex RNA analytes [31, 32]

In this study, we introduce a novel dual-guide CRISPR/Cas13 platform, in which the conventional single guide RNA is divided into two separate RNA fragments connected through engineered 5- or 7-nucleotide complementary linker sequences. This innovative design preserves the catalytic activity of LwaCas13a at levels comparable to the conventional single-guide system, while substantially enhancing its specificity. We demonstrate that spatially separating the guide RNA reduces off-target binding, which is a major limitation of current Cas13-based diagnostics. We validate the improved specificity and functionality of this dual-guide system through two clinically relevant applications: the sensitive and specific detection of SARS-CoV-2 RNA, demonstrating its potential for pathogen diagnostics, and the identification of single-nucleotide variants (SNVs) in the oncogene KRAS, including the differentiation between wild-type and key mutant alleles such as G12D and G12C, demonstrating its potential for cancer diagnosis. Our results highlight the robustness and versatility of the dual-guide approach, providing an innovative platform that addresses key challenges in nucleic acid diagnostics, with particular relevance for precision medicine.

## MATERIAL AND METHODS

### Patient samples

For the SARS-CoV-2 analysis, we used five saliva control samples from healthy adult volunteers and ten saliva samples from individuals with confirmed SARS-CoV-2 infection by RT-qPCR. All samples were provided by the Biobank of the Public Health System of Andalusia (reference S2000262), and informed consent was obtained from all participants. Total RNA was extracted from each sample using the Qiagen RNeasy Mini Kit (Qiagen, Hilden, Germany) according to the manufacturer’s instructions. Purified RNA samples were stored at –80 °C until further use to preserve integrity for downstream analyses.

### Cell lines and Culture media

MiaPaca-2 (CRL-1420, ATCC) were maintained in DMEM medium (Dulbecco’s modified Eagle medium high glucose) (Biowest, Nuaillé, France) supplemented with 10% foetal bovine serum (FBS) (Biowest) and 1% penicillin/streptomycin (Biowest). BxPC3 (CRL-1687) and AsPC1 (CRL-1682, ATCC) cells were maintained in RPMI-1640 medium (Biowest), supplemented with 10% FBS and 1% penicillin/streptomycin. Cells were maintained in incubators at 37°C with 5% CO_2_ atmosphere and 85%–90% relative humidity. Absence of *Mycoplasma spp*. in cultured cells was routinely tested by a PCR-based assay (Minerva Biolabs, Skillman, NJ, USA).

### RT-qPCR

To confirm SARS-CoV-2 positivity in patient samples, total RNA was reverse-transcribed into cDNA using the High-Capacity cDNA Reverse Transcription Kit (Applied Biosystems, Waltham, MA, USA) with RNase Inhibitor (Applied Biosystems), following the manufacturer’s instructions. Quantitative PCR (qPCR) was then performed using Fast SYBR™ Green Master Mix (Applied Biosystems) on a QuantStudio™ 6 Flex Real-Time PCR System (96-well format; Applied Biosystems). The amplification protocol was as follows: 95 °C for 5 s; followed by 40 cycles of 96.5 °C for 10 s and 62–64 °C for 30 s; and a melt curve step consisting of 95.6 °C for 10 s, 55 °C with a ramp rate of 0.05 °C/s, and 95 °C for 30 s. All reactions were performed in duplicate using the primers listed in **Supplementary Table S1**.

### RNA synthesis

To generate synthetic RNA targets, SARS-CoV-2 cDNA was PCR-amplified using KAPA HiFi HotStart ReadyMix (Kapa Biosystems, Wilmington, MA, USA) and primers listed in **Supplementary Table S1**. The resulting double-stranded DNA (dsDNA) amplicons were gel-purified using the MinElute Gel Extraction Kit (Qiagen). *In vitro* transcription was performed by incubating the purified dsDNA overnight at 37 °C with T7 RNA polymerase (New England Biolabs, Ipswich, MA, USA) in the presence of RNase inhibitor (Applied Biosystems) and rNTPs (New England Biolabs). The transcribed RNA was purified using the MEGAclear Transcription Clean-Up Kit (Applied Biosystems) according to the manufacturer’s instructions.

### Electrophoretic Mobility Shift Assay (EMSA)

RNA–protein interactions were assessed using an electrophoretic mobility shift assay (EMSA) optimized for Cas13-based complexes. First, dual guide RNA complexes were prepared by annealing 10 µL of dcrRNA (10 µM, Integrated DNA Technologies, Coralville, IA, USA) with 10 µL of dtracrRNA (10 µM, Integrated DNA Technologies) in the presence of 5 µL of annealing buffer (Synthego, Menlo Park, CA, USA). This mixture was incubated in a thermocycler using the following program: 95 °C for 4 minutes, 65 °C for 5 minutes, 25 °C for 5 minutes, and then held at 4 °C. Recombinant LwaCas13a (57 µM, GenScript, Piscataway, NJ, USA) was diluted in 2 buffer (New England Biolabs, Ipswich, MA, USA) supplemented with BSA to a working concentration of 4 µM. To assemble the Cas13-RNP complex (2 µM), the diluted Cas13 was mixed with the annealed crRNA:tracrRNA duplex and incubated for 10 minutes at room temperature. Samples were then kept on ice until further use. Binding reactions were prepared by combining appropriate volumes of the Cas13-RNP complex with binding buffer (20 mM HEPES, pH 7.5, 250 mM KCl, 2 mM MgCl₂, 0.01% Triton X-100, 10% glycerol; all reagents from Sigma-Aldrich, St. Louis, MO, USA) to reach final complex concentrations of 0, 10, 100, or 1000 nM. Subsequently, 1 µL of target RNA (100 ng/µL) previously synthesized was added to each reaction, followed by incubation for 1 hour at 37 °C. Following incubation, each reaction was loaded onto a precast native 4–20% polyacrylamide gel (Bio-Rad, Hercules, CA, USA). Electrophoresis was performed in 1× TBE buffer supplemented with 2 mM MgCl₂ at 100 V for 75 minutes at 4 °C. After electrophoresis, the gel was stained in 1× TBE containing GelRed® nucleic acid stain (Biotium, Fremont, CA, USA) and 2 mM MgCl₂. Visualization of RNA–protein complexes was performed using a Gel Doc™ EZ Imager system (Bio-Rad). Images were analysed and band intensities quantified using ImageJ software version 1.53q (NIH, Bethesda, MD, USA).

### Cas13 Collateral Cleavage Detection Assay (SHERLOCK)

Detection of target RNA or DNA sequences was performed using a two-step SHERLOCK (Specific High-Sensitivity Enzymatic Reporter Unlocking) assay optimized for LwaCas13a collateral activity using both single- and dual-guide CRISPR systems.

(1) Reverse Transcription and PCR Amplification. Total RNA extracted from patient samples (including both SARS-CoV-2 positive and negative cases) was reverse transcribed into complementary DNA (cDNA) using the High-Capacity cDNA Reverse Transcription Kit with RNase Inhibitor (Applied Biosystems), following the manufacturer’s instructions. For SARS-CoV-2 samples, known amounts of cDNA (10 ng or 1 ng per reaction) were used. In the case of KRAS detection, genomic DNA (gDNA) extracted with using a QIAamp DNA mini kit (Qiagen) following the manufacturer’s instructions, instead at concentrations of 10, 1, or 0.1 ng per reaction. Target sequences were amplified via endpoint PCR using gene-specific primers (Table X) and GoTaq® G2 DNA Polymerase Master Mix, colorless 2× (Promega, Madison, WI, USA) followed by incubation in a thermocycler with the following program: 95 °C for 4 min, 65 °C for 5 min, 25 °C for 5 min, and held at 4 °C.
(2) *In Vitro* Transcription and Cas13 Detection. Cas13-based detection was carried out in 384-well microplates. The plate reader (NanoQuant, Tecan, Männedorf, Switzerland) was preheated to 37 °C, and all reagents were maintained on ice until transferred to the plate. LwaCas13a (GenScript) was diluted to a final concentration of 450 nM in Buffer 2 (New England Biolabs) supplemented with BSA. The detection mix (19 µL per well) was assembled in the following order: RNase-free water, HEPES buffer (pH 6.8, final concentration 20 mM; Sigma), guide RNA (45 nM; either a synthetic sgRNA from Integrated DNA Technologies, or a dual-guide complex). The dual-guide complex was previously formed by annealing 10 µL of dcrRNA (10 µM, IDT) with 10 µL of dtracrRNA (10 µM, IDT) in the presence of 5 µL of annealing buffer (Synthego), followed by incubation in a thermocycler with the following program: 95 °C for 4 min, 65 °C for 5 min, 25 °C for 5 min, and held at 4 °C. The mix was completed by adding LwaCas13a protein (final 45 nM), murine RNase inhibitor (2 U/µL; New England Biolabs), rNTP solution (1 mM each; New England Biolabs), T7 RNA polymerase (0.125 U/µL; New England Biolabs), and the RNaseAlert substrate (125 nM; IDT). The reaction was initiated by adding 1 µL of the PCR product to each well. Fluorescence was monitored every 5 minutes for 3 hours at 37 °C using excitation and emission wavelengths of 485 nm and 520 nm, respectively. All reactions were performed in triplicate and protected from light throughout the procedure to preserve reporter integrity.

### AlphaFold 3 structure prediction and analysis

Structural modelling of the Cas13a protein complex with either the conventional single-guide RNA (sgRNA) or dual-guide RNA constructs was performed using AlphaFold 3 (DeepMind Technologies Ltd., London, UK, and Isomorphic Labs Ltd., London, UK) via the AlphaFold Server server https://alphafoldserver.com/ in (April, 2025) [33]. The LwaCas13a amino acid sequence was obtained from UniProt (ID: U2PWF1), due to the absence of an experimentally resolved structure. Input RNA sequences corresponded to either the canonical sgRNA or the dual-guide RNA constructs. For each complex, the five ranked models generated by AlphaFold were analysed, and the top-ranked model (Rank 1) was selected for further study. These models were subsequently imported into UCSF ChimeraX (version 1.2.5) for visualization and detailed structural examination. The B-factor attribute was employed to assess regional variability among models.

### Statistical Analysis

Statistical analyses on data obtained were performed and represented with the GraphPad Prism software (version 8.0.1) (GraphPad Software, La Jolla, CA, USA). All data are presented as mean ± SEM. Two-way ANOVA was used for multiple comparisons, followed by appropriate post hoc tests. Statistical significance is indicated as follows: * p < 0.05, ** p < 0.01, *** p < 0.001, **** p < 0.0001). The specific tests used are detailed in each figure legend.

### Figures creation

For formatting the figures Adobe Photoshop version 13.0.1 (Adobe, San José, CA, USA) was used. Figure 1A was designed using BioRender (BioRender.com). This design served as the base for the subsequent creation of Figures 2A, 3A and Graphical abstract, which were then adapted and finalized using Adobe Photoshop (Adobe).

**Figure 1.**
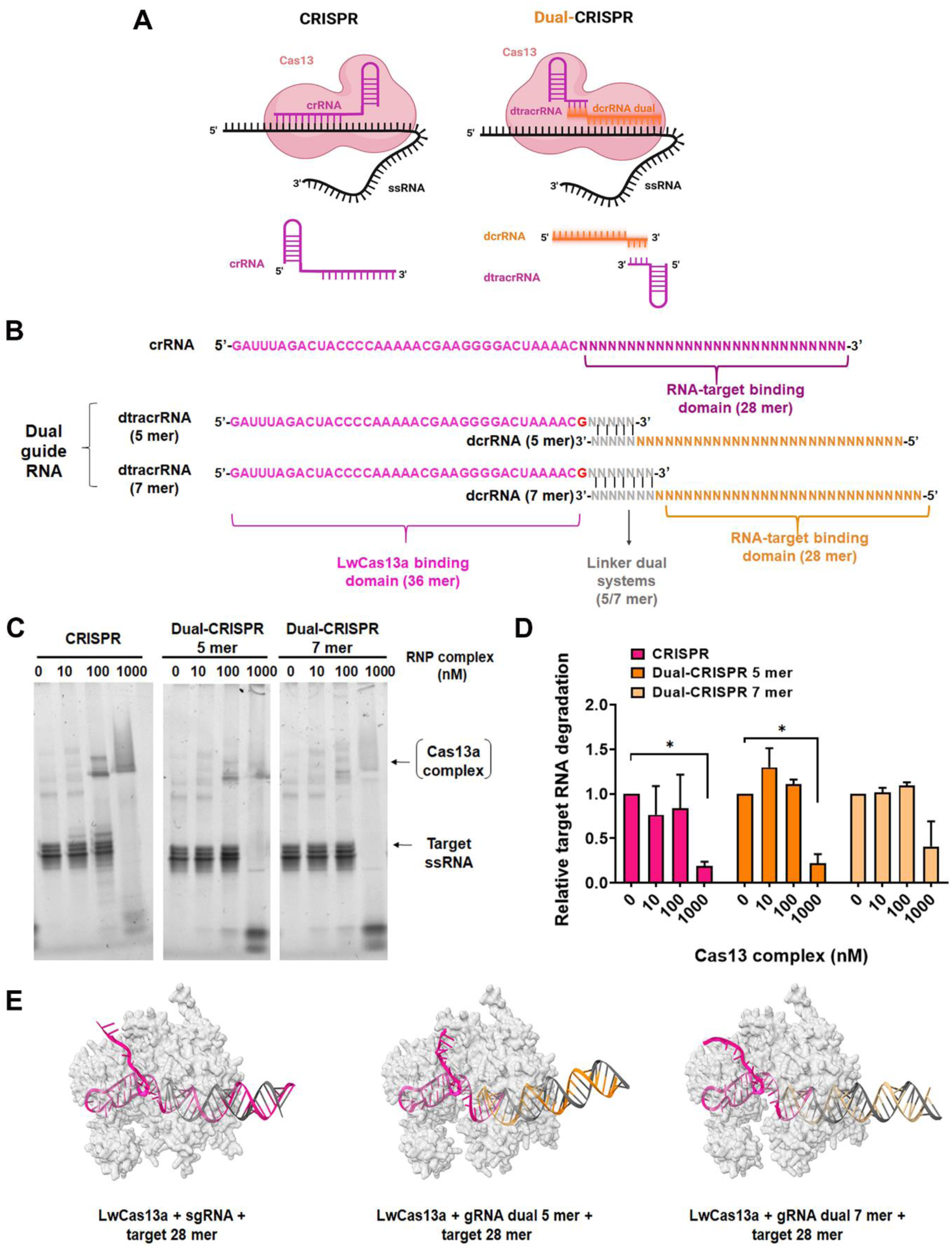
Designing and characterization of the novel Dual-CRISPR system for Cas13. A) Schematic representation of conventional CRISPR and the newly developed Dual-CRISPR architecture for Cas13. In the conventional system, Cas13 binds to a single crRNA molecule that acts as both the Cas-binding and target-recognition domain. In the Dual-CRISPR system, these functions are dissociated into two distinct RNAs: a variable dcrRNA responsible for target binding, and a constant dtracrRNA that mediates interaction with Cas13 and dcrRNA. The final complex comprises three components: LwaCas13a protein, the variable dcrRNA targeting the sequence of interest, and the constant dtracrRNA. Created in BioRender. Aguilar, A. (2026) https://BioRender.com/gxqr13o. B) Sequences of the CRISPR and Dual-CRISPR (5-mer and 7-mer linkers) components used with LwaCas13a. The Cas13-binding domain is highlighted in pink, the 28-nucleotide spacer targeting the viral RNA is highlighted in purple (Conventional system) or orange (Dual-system), and the linker sequences for the Dual-CRISPR constructs are shown in gray. C) Electrophoretic mobility shift assay (EMSA) of CRISPR and Dual-CRISPR (5-mer and 7-mer) complexes incubated with their SARS-CoV-2 RNA target. Complexes were assembled *in vitro* and incubated with increasing concentrations (0, 10, 100, and 1000 nM) of the CRISPR/Cas13 complexes. D) Quantification of residual target RNA (ssRNA) from EMSA in panel C. Band intensities corresponding to the free RNA target were quantified and normalized to the 0 nM RNP complex condition (no Cas13a). A decrease in intensity reflects increasing target RNA degradation due to Cas13a activity. Bars represent mean ± SEM of n = 3 separate experiments. E) Predicted 3D structures of the CRISPR and Dual-CRISPR (5-mer and 7-mer) complexes bound to the target RNA (28 nt), modelled using AlphaFold 3.

**Figure 2.**
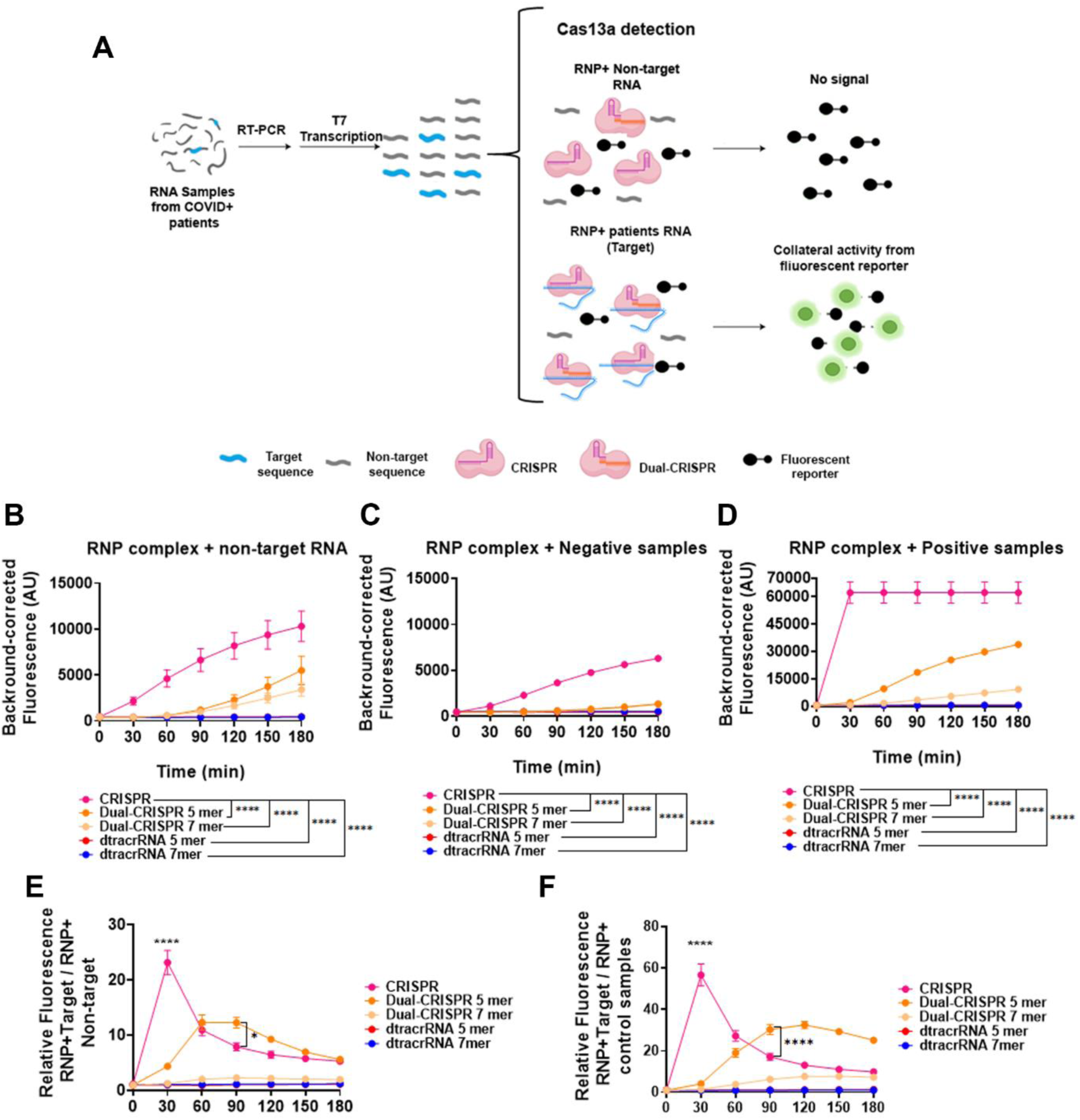
Evaluation of the functionality of the Dual-CRISPR/Cas13 system for SARS-CoV-2 virus detection. A) Schematic of the PCR-adapted-SHERLOCK adapted method for SARS-CoV-2 virus detection for CRISPR and Dual-CRISPR system. B-D) Corrected fluorescence of RNP-CRISPR, dual-CRISPR 5mer and 7mer complexes with non-target RNA (RNA from HEK293T cells) (B), with negative samples (healthy controls) (C) and with positive samples (SARS-CoV-2 RNA target) (D) after incubation with the fluorescent reporter for 3 hours. E) Relative fluorescence of RNP complexes with RNA from positive samples (target RNA) versus fluorescence of RNP complexes with non-target RNA (HEK293T). F) Relative fluorescence of RNP complexes with RNA from positive samples (target RNA) versus fluorescence of RNP complexes with negative samples. Statistical analysis: 2-way ANOVA (Dunnett’s multiple comparison test) of mean fluorescence at all times (B-D) and at specific times such as 30 min and 90 min (E, F) (* p < 0.05, ** p < 0.01, *** p < 0.001, **** p < 0.001). Values represent the mean ± SEM of 3 separate experiments.

## RESULTS

### Design and structural characterization of the Dual-CRISPR system for LwaCas13a

To address the limitations of conventional single-guide designs in CRISPR/Cas13 systems, we engineered a modular dual-guide architecture, coined Dual-CRISPR. Unlike the conventional CRISPR system where a single crRNA molecule mediates both Cas13 binding and target recognition, our Dual-CRISPR system dissociates these functions into two distinct components: a variable dual crRNA (dcrRNA), which contains the spacer sequence complementary to the RNA target, and a constant dual tracrRNA (dtracrRNA), which mediates interaction with LwaCas13a and the dcrRNA. These two components interact via short complementary linker sequences (5 or 7 nucleotides), de novo designed in this study to enable stable hybridization and efficient complex formation (Figure 1A).

For the initial characterization of the system, we designed guide RNAs targeting a region within the spike (S) gene-derived RNA of SARS-CoV-2. The structure of the single-guide and dual-guide constructs are shown in Figure 1B, with the conserved Cas13-binding domain, 28-nt spacer, and our custom-designed linker regions highlighted in different colours. The full sequences of all guide RNAs used in this study are listed in **Supplementary Table S2**.

Complex formation and RNA binding capacity were evaluated by electrophoretic mobility shift assays (EMSA), (Figure 1C). Both dual-guide constructs (5-mer and 7-mer) efficiently formed RNP complexes with the SARS-CoV-2 target RNA, showing similar or higher RNA binding capacity compared to the conventional Cas13 system. In all conditions, increasing concentrations of the RNP complex led to progressive degradation of the target ssRNA, as evidenced by the decrease in free RNA signal. Quantification of band intensities (Figure 1D) confirmed a concentration-dependent enzymatic activity, consistent with efficient target cleavage by both dual-guide and conventional Cas13 constructs.

To further investigate the integrity of our dual-CRISPR design, 3D structural models were generated using, the latest generation of AI-driven molecular modeling system, AlphaFold 3 [33]. These models accurately predict the conformations of LwaCas13a bound to both single-guide (sgRNA) and dual-guide RNA constructs, alongside the 28-nucleotide target RNA. The dual-guide models, particularly the 5-mer linker variant, exhibited overall structural similarity to the conventional CRISPR/Cas13 complex, suggesting that the introduction of a split guide does not disrupt the formation of a functional ribonucleoprotein complex (Figure 1E). A more detailed analysis of these models, including structural alignment and conformational features, is provided in the **Supplementary Figure S1**.

### Validation of the Dual-CRISPR/Cas13 system for RNA detection with a SHERLOCK-based assay

In order to evaluate the diagnostic potential of the Dual-CRISPR/Cas13 system, a SHERLOCK-based assay for SARS-CoV-2 RNA was performed. All samples were previously validated by RT-qPCR to confirm their SARS-CoV-2 status (positive or negative) **(Supplementary Figure S2)**. Traditional SHERLOCK protocols rely on isothermal amplification of the target sequence, typically using recombinase polymerase amplification (RPA) followed by *in vitro* transcription to generate RNA for Cas13 detection [34]. However, while RPA is a valuable amplification method, under our experimental conditions, it exhibited a higher susceptibility to non-specific amplification and cross-contamination. Several primer pairs targeting the S gene region of SARS-CoV-2 were tested, but false-positive signals were detected in negative controls **(Supplementary Figure S3)**. In light of the limited specificity and increased contamination risk associated with RPA, under these experimental conditions, we opted to replace this step with PCR amplification.

For this approach, PCR primers were designed to flank the target region within the SARS-CoV-2 spike (S) gene, with a T7 promoter sequence incorporated into the forward primer **(Supplementary Table S1)**. This allowed the resulting amplicons to act as templates for *in vitro* transcription, producing RNA for subsequent detection by Cas13. A schematic representation of the adapted assay workflow is shown in Figure 2A. PCR amplicons containing a T7 promoter are incubated in a single reaction with T7 RNA polymerase, Cas13 RNPs, and a fluorescent RNA reporter. During incubation, the DNA amplicons are transcribed into RNA. Upon specific recognition of the RNA target, Cas13 is activated and cleaves the fluorescent reporter via its collateral (*trans*) activity, generating a measurable fluorescence signal [17].

The predicted secondary structures of the *in vitro* transcribed RNA targets used in these assays is shown in **Supplementary Figure S4A**. These structural models, generated using FoRNA software, reveal distinct conformations that may influence target accessibility and Cas13-mediated cleavage.

We compared the collateral activity of standard CRISPR/Cas13 and Dual-CRISPR/Cas13 systems (5-mer and 7-mer) using non-target RNA extracted from HEK293T cells, RNA from healthy donors (SARS-CoV-2 negative samples confirmed by RT-qPCR), and target RNA from SARS-CoV-2 positive patients confirmed by RT-qPCR. Fluorescence signal resulting from reporter cleavage was monitored over a 3-hour period. Both Dual-CRISPR variants generated substantially lower fluorescence signals in the presence of non-target RNA and negative samples (healthy control) compared to the conventional system (Figure 2B-C), indicating improved specificity. In contrast, RNA from positive samples (particularly when tested with the 5-mer variant) yielded markedly higher fluorescence, reaching levels comparable to the standard CRISPR complex. Although standard CRISPR complex had faster kinetic, the dual system achieved comparable levels after 180min (Figure 2D). These results demonstrate the diagnostic potential and functionality of the Dual-CRISPR system.

Although absolute fluorescence was lower for Dual-CRISPR, target discrimination by normalizing signals from patient samples (target) against both negative controls (Figures 2E**– F)**. The Dual-CRISPR 5-mer variant demonstrated robust target-specific activation with improved specificity, particularly during longer incubation periods (>60 min), a timeframe in which the conventional CRISPR/Cas13 system showed a tendency toward increased background noise under our experimental conditions.

### Evaluation of mismatch sensitivity in Dual-CRISPR vs. CRISPR for SARS-CoV-2 detection

The inherent tolerance of conventional Cas13 systems to single-nucleotide mismatches presents a significant challenge for their application in molecular diagnostics, often leading to reduced specificity and potential false positives, particularly when differentiating closely related targets or identifying single-nucleotide variants (SNVs). To evaluate mismatch sensitivity, we further explored the specificity of the Dual-CRISPR/Cas13 system by assessing its ability to discriminate single and double mismatches in the guide-target pairing. We focused on the 5-mer dual-guide, which had previously demonstrated the most favourable performance in terms of target recognition and signal specificity. To this end, we designed several guides RNA containing either one (OFF5) or two (OFF3&5) synthetic mismatches relative to the SARS-CoV-2 target sequence. Positions 5 and 3&5 within the guide sequence were selected based on previous studies which identified these sites as having the greatest destabilizing effect on guide-target pairing [17, 35] (Figure 3A).

**Figure 3.**
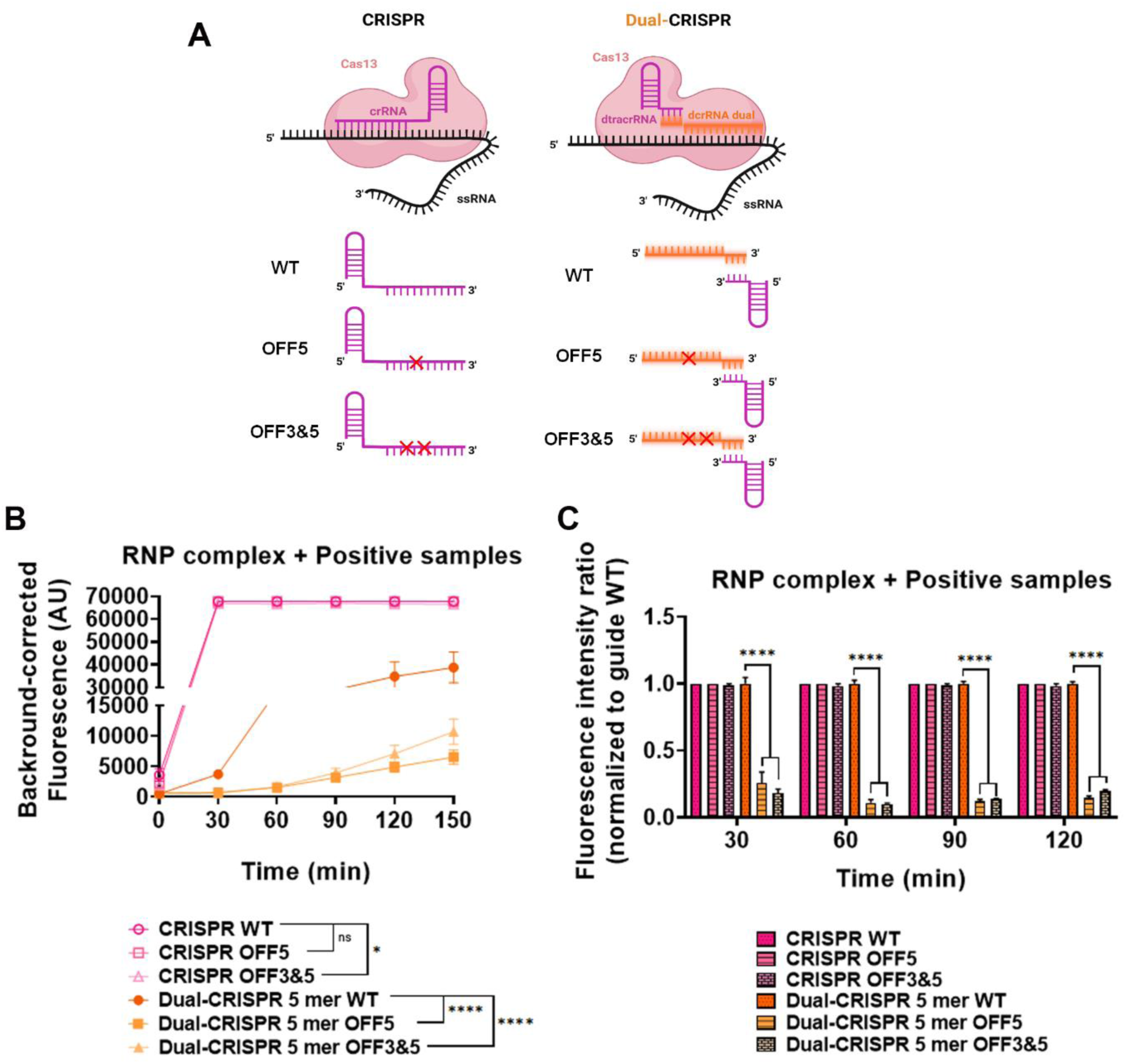
Evaluation of the specificity of Dual-CRISPR/Cas13 vs. CRISPR for SARS-CoV-2 virus detection. A) Scheme of the different guides designed for CRISPR and dual-CRISPR 5mer. The WT guide binds perfectly to the SARS-CoV-2 target sequence. The OFF5 and OFF3&5 guides have 1 and 2 mismatches with the SARS-CoV-2 target sequence, respectively. B) Corrected fluorescence of RNP-CRISPR and dual-CRISPR 5mer complexes with RNA from patient samples (target RNA) after incubation with the fluorescent reporter for 150 minutes. C) Fluorescence intensity ratio normalized to the WT guide (CRISPR and Dual-CRISPR 5 mer, respectively) at different time points (30, 60, 90 and 120 minutes). Statistical analysis: 2-way ANOVA with a Dunnett’s multiple comparisons test was used to compared mean fluorescence at all times (B) and at specific times (C) comparing between WT and different mismatches) (* p < 0.05, ** p < 0.01, *** p < 0.001, **** p < 0.001). Values represent the mean ± SEM of 3 separate experiments.

Then, the collateral activity of Cas13 was compared with either wild-type or mismatched guides using the RNA extracted from SARS-CoV-2 positive samples as target. Fluorescence generated by reporter cleavage was monitored in both the conventional CRISPR and Dual-CRISPR systems. The standard CRISPR system exhibited minimal variation in fluorescence signals between wild-type and altered guides (OFF5 and OFF3&5), even with two mismatches, indicating a relatively high tolerance for sequence divergence (Figure 3B).

These results are in line with the previous studies mentioned above, which reported that single mismatches do not substantially affect Cas13 activity [23, 36]. In contrast, the Dual-CRISPR 5-mer exhibited a substantial decrease in signal intensity, reducing fluorescence by 80-85% approximately, when using mismatched guides, with just one mismatch causing a significant reduction in fluorescence (Figure 3B**, orange dots vs. orange squares and triangles)**. To better quantify the impact of mismatches on target recognition, fluorescence signals were normalized to the corresponding wild-type guide signal at different time points. While the conventional CRISPR system has been widely used and provides reliable results, the Dual-CRISPR system demonstrated notably enhanced discrimination against single and double mismatches (Figure 3C). This improvement represents an important advancement toward increasing diagnostic accuracy.

### Improved Specificity of Dual-CRISPR-Cas13 System in Detecting Oncogenic KRAS Variants

Next, the efficiency of Dual-CRISPR-Cas13 system in distinguishing clinically relevant single-nucleotide variants (SNVs) was evaluated. Given the relevance of detecting oncogenic mutations with high specificity for precise patient stratification and effective treatment decisions, the next phase of this research focuses on evaluating the implementation of this technology in these settings. Detecting oncogenic mutations with high specificity is critical in clinical diagnostics for accurate patient stratification and treatment guidance. Single-nucleotide variants (SNVs) in the KRAS gene, such as G12D and G12C, are prevalent in various cancers, including pancreatic adenocarcinoma, and their precise identification is crucial for targeted therapies such as monoclonal antibody-based therapies. Conventional Cas13 systems, while sensitive, often struggle to reliably discriminate between such closely related SNVs within a complex genomic background, potentially leading to misdiagnosis. We thus evaluated the specificity of our Dual-CRISPR-Cas13 system for detecting these clinically relevant KRAS mutations (G12D and G12C) in a cell-based genomic DNA model. The presence of these specific KRAS was validated in different pancreatic cell lines **(Supplementary Figure S5)**, ensuring a reliable model for specificity testing. Genomic DNA was thus extracted from well-characterized pancreatic cell lines: BxPC-3 (KRAS WT), AsPC-1 (KRAS G12D), and MIA-PaCa2 (KRAS G12C), each homozygous for their respective alleles. The predicted secondary structures of the *in vitro* transcribed RNA targets used for KRAS are shown in **Supplementary Figure S4B-D**. A guide RNA specific for the G12D mutation was used in both the conventional CRISPR/Cas13 assay and our Dual-CRISPR/Cas13 configuration [35]. This design allowed for the direct comparison of their abilities to selectively detect the KRAS G12D allele, while effectively discriminating against the wild-type sequence (representing a single mismatch) and the KRAS G12C sequence (representing two mismatches) (Figure 4A). To enable detection from genomic DNA, the SHERLOCK workflow was adapted from the RNA-based format previously used for SARS-CoV-2 (Figure 2A) to use genomic DNA as input for KRAS mutation analysis.

**Figure 4.**
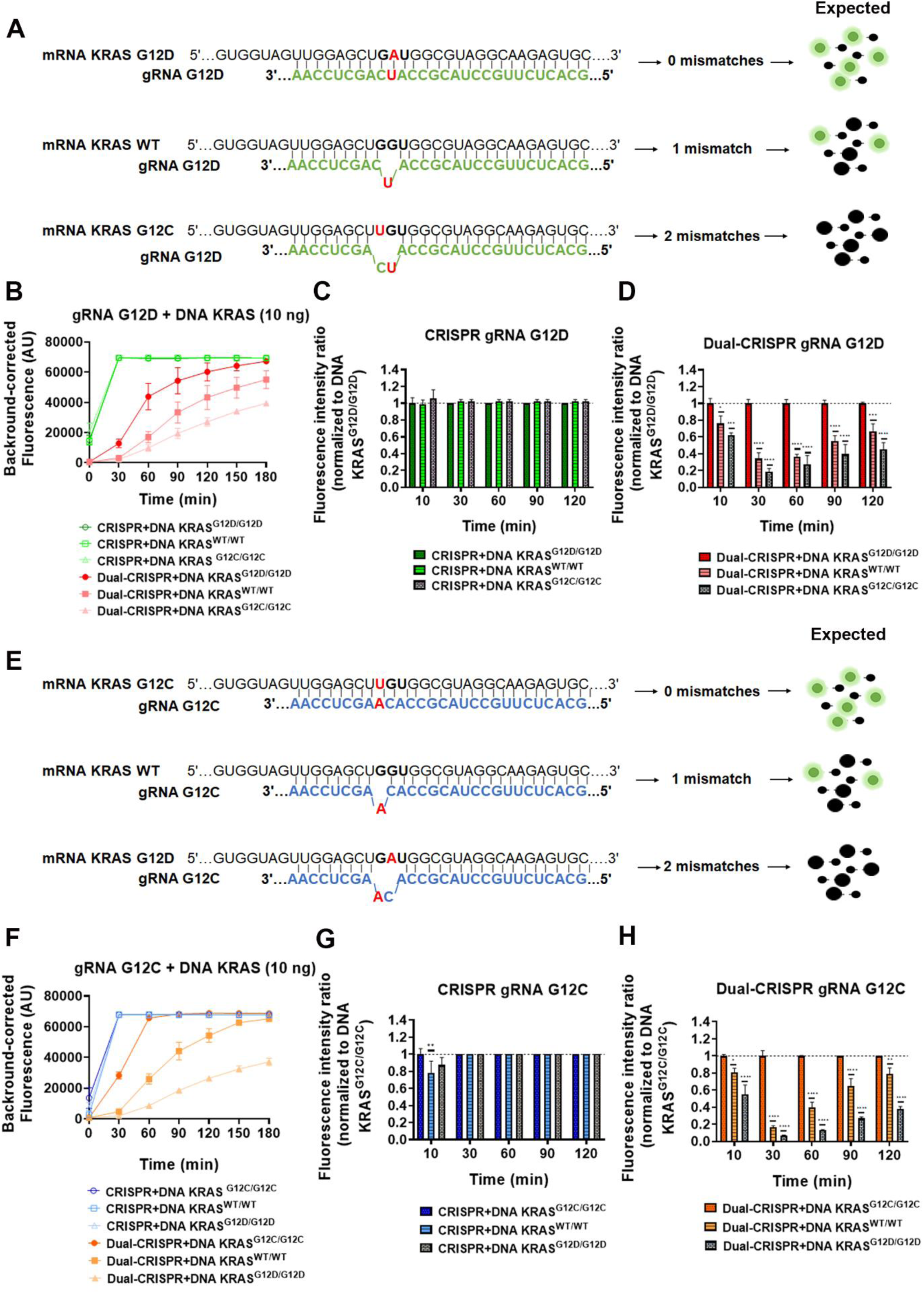
Evaluation of CRISPR and Dual-CRISPR Specificity in the Detection of KRAS G12D and G12C Variants. A) Schematic representation of the KRAS G12D guide, which is designed for use with CRISPR and Dual-CRISPR systems, binding to different RNA targets (KRAS G12D, KRAS WT or KRAS G12C). The expected output from the SHERLOCK system is shown based on the number of mismatches. The KRAS G12D guide binds perfectly to the KRAS G12D target sequence. The KRAS WT and KRAS G12C targets have one and two mismatches with the G12D guide, respectively. B) Corrected fluorescence of RNP-G12D guides (CRISPR and Dual-CRISPR) with DNA from pancreatic cell lines harbouring G12D, WT or G12C alleles (10 ng), after 180 minutes’ incubation with the fluorescent reporter. C) Fluorescence intensity ratio normalised to KRAS G12D DNA (10 ng) using RNP-G12D guides (CRISPR) at different time points (30, 60, 90 and 120 minutes). D) Fluorescence intensity ratio normalised to KRAS G12D DNA (10 ng) using RNP-G12D guides (Dual-CRISPR) at different time points (30, 60, 90 and 120 minutes). E) A schematic representation of the binding of the KRAS G12C guide, which is designed for the CRISPR and Dual-CRISPR systems, to different RNA targets (KRAS G12C, KRAS WT or KRAS G12D) and the expected SHERLOCK output, based on the number of mismatches. The KRAS G12C guide binds perfectly to the KRAS G12C target sequence. The KRAS WT and KRAS G12D targets have one and two mismatches with the G12C guide, respectively. F) Corrected fluorescence of RNP-G12C guides (CRISPR and Dual-CRISPR) with DNA from pancreatic cell lines harbouring G12C, WT or G12D alleles (10 ng) after 180 minutes of incubation. G) Fluorescence intensity ratio normalised to KRAS G12C DNA (10 ng) using RNP-G12C guides (CRISPR) at different time points (30, 60, 90 and 120 minutes). H) Fluorescence intensity ratio normalised to KRAS G12C DNA (10 ng) using RNP-G12C guides (Dual-CRISPR) at different time points (30, 60, 90 and 120 minutes). Statistical analysis: 2-way ANOVA with a Dunnett’s multiple comparisons test was used to compare specific targets with non-specific sequences (1 and 2 mismatches) at each time point (* p < 0.05, ** p < 0.01, *** p < 0.001, **** p < 0.0001). Data are presented as the mean ± SEM from at least three independent experiments.

We then compared the collateral activity of G12D-gRNA:Cas13 complexes across different DNA targets (G12D, WT, and G12C) using genomic DNA (10 ng). The conventional CRISPR/Cas13 system showed minimal signal profile differences over time between the specific G12D and non-specific WT and G12C targets (Figure 4B-C). In contrast, our Dual-CRISPR/Cas13 configuration consistently demonstrated superior mismatch discrimination exhibiting higher specificity by clearly distinguishing the G12D target from WT (1 mismatch) and G12C (2 mismatches) sequences (Figure 4B and D). This enhanced specificity was maintained at lower DNA inputs (1 ng and 0.1 ng; **Supplementary Figures S6A–C and S7A– C**). At lower inputs (0.01 ng and 0.001 ng), no signal was detected for any target, even to conventional CRISPR system, indicating a detection threshold of approximately 0.1 ng under current assay conditions **(Supplementary Figure S8)**.

Using a G12C-specific guide RNA, both systems were evaluated for their ability to discriminate the G12C mutation. The Dual-CRISPR configuration consistently and clearly distinguished G12C from WT and G12D sequences (Figure 4F and H), while the conventional system failed to differentiate between specific and non-specific targets (Figure 4F-G). Notably, the high specificity of the Dual-CRISPR approach remain robust across all tested DNA concentrations **(Supplementary Figures S6D–F and S7D–F)**.

These findings highlight the superior mismatch discrimination and enhanced diagnostic accuracy achieved with the Dual-CRISPR/Cas13 system relative to the conventional CRISPR assay.

As summarized in **Table 1**, our Dual-CRISPR/Cas13 system demonstrates a markedly improved ability to discriminate between clinically relevant KRAS single-nucleotide variants (SNVs), while avoiding false-positive signals from closely related sequences that the conventional assay fails to distinguish.

**Table 1.** Comparative analysis of mismatch discrimination between the standard and Dual-CRISPR/Cas13 systems targeting KRAS single-nucleotide variants (SNVs). Each guide RNA was tested against matched (on-target) and mismatched sequences (1 or 2 nucleotide differences). *In the Standard CRISPR-Cas13 system, guide RNA refers to single-guide RNA (sgRNA) that are designed to be specific for the target (either G12D or G12C). In the Dual-CRISPR system, the specific guide RNA refers to the variable dual crRNA (dcrRNA), which contains the sequence complementary to the target. In bold and *italic*, the correct calls.

| Guide RNA* | Target DNA Tested | Standard CRISPR-Cas13 | Dual-CRISPR-Cas13 |
| --- | --- | --- | --- |
| G12D-specific | KRAS G12D (On-Target) | **Detected** | **Detected** |
| G12D-specific | KRAS WT (1 Mismatch) | Detected (False Positive) | **Not Detected (Accurate)** |
| G12D-specific | KRAS G12C (2 Mismatches) | Detected (False Positive) | **Not Detected (Accurate)** |
| G12C-specific | KRAS G12C (On-Target) | **Detected** | **Detected** |
| G12C-specific | KRAS WT (1 Mismatch) | Detected (False Positive) | **Not Detected (Accurate)** |
| G12C-specific | KRAS G12D (2 Mismatches) | Detected (False Positive) | ***Not Detected (Accurate)*** |

## DISCUSSION

In this study, we developed and validated a dual-guide RNA architecture for CRISPR-Cas13a that enhances target specificity while maintaining robust RNA cleavage activity. By splitting the conventional single guide RNA into two distinct components (dcrRNA and dtracrRNA) linked via complementary sequences, the system introduces an additional layer of molecular recognition and enhances mismatch discrimination. The platform was successfully applied to detect both SARS-CoV-2 RNA and oncogenic KRAS mutations, demonstrating versatility across clinically relevant applications.

In the context of viral diagnostics, the dual-guide system effectively identified SARS-CoV-2 RNA with high specificity, particularly the 5-mer variant, even at long incubation times. While its sensitivity was slightly lower than the conventional single-guide Cas13 system at early time points, the dual architecture provided greater signal stability and significantly reduced background noise. Moreover, the dual-guide system presented a pronounced and significant decrease in signal intensity when challenged with guide RNAs containing single or double synthetic mismatches relative to the target sequence (Figure 3B, C). This is in great contrast to the conventional CRISPR system, which showed minimal differentiation in fluorescence signals between wild-type and mismatched guides (Figure 3B, C). The strategic selection of mismatch positions (5 and 3&5) based on prior literature further underscores the deliberate approach taken to challenge the system’s discriminatory power at sites known to have a significant impact on guide-target pairing.

Previous CRISPR-based diagnostics using Cas13a, such as SHERLOCK [17, 37], or Cas13-based RPA amplification platforms [38], can achieve sensitive and multiplexed detection of viral RNA. However, despite their speed and simplicity, these approaches are often susceptible to nonspecific amplification, carryover contamination, and amplification artefacts, particularly in resource-limited settings. In contrast, our Dual-CRISPR platform was integrated with a PCR-based pre-amplification strategy, providing a more controlled and robust workflow that reduces nonspecific background and improves assay reproducibility.

Beyond pathogen detection, we demonstrated that the dual-guide CRISPR-Cas13 system is also effective in distinguishing clinically relevant single-nucleotide variants (SNVs), using KRAS mutations as a model. Specifically, we showed that our system could robustly discriminate between the oncogenic G12D and G12C alleles and the wild-type KRAS sequence, achieving improved mismatch sensitivity over the conventional single-guide approach (Figure 4B, D, F, H). The standard CRISPR system, under the same conditions, failed to clearly differentiate between specific and non-specific targets, exhibiting significant overlap in signal profiles (Figure 4B, C, F, G). This superior mismatch discrimination was maintained even at lower genomic DNA inputs with a detection threshold of approximately 0.1 ng under current assay conditions, highlighting the potential of the Dual-CRISPR system for applications where sample availability or target concentration is limited (Figures S6 and S7).

This distinction is critical, as accurate identification of KRAS mutations has direct implications for the diagnosis, prognosis, and targeted treatment of various cancers, including pancreatic, colorectal, and non-small lung carcinomas [39–41]. Our system’s ability to clearly distinguish between single-base changes represents a critical step forward in precision oncology diagnostics.

Previous studies using Cas13 for SNV detection have relied on guide engineering or tuning buffer conditions, often with limited success in complex backgrounds [21, 23]. However, such methods still require extensive optimization of guide-target interactions or reliance on engineered Cas13 variants, and remain challenging for broadly applicable diagnostic assays. Using this aproach, by requiring dual guide-target recognition, false positives are substantially reduced, making the approach suitable for liquid biopsy applications where mutant alleles may be present at low frequencies.

Several studies have recently reported that split-guide strategies in CRISPR-Cas12 could improve single-nucleotide mismatch discrimination when detecting microRNAs [27, 30]. These systems are based on the same principle of spatial separation between RNA domains to increase target specificity, a strategy conceptually similar to our dual-guide design for Cas13. Notably, these split-Cas12 approaches have been validated primarily in short RNA targets, such as microRNAs (<22 nt), and rely on high concentrations of protein and RNA to maintain detectable activity. In contrast, our dual-guide Cas13 system extends the applicability of the split-guide concept to longer and more complex RNA targets, including full-length viral genomes (SARS-CoV-2) and endogenous human mRNAs containing clinically relevant SNVs (e.g., KRAS mutations). This broader target range, combined with the robust discrimination of closely related sequences, highlights the versatility and diagnostic potential of the dual-guide Cas13 platform.

Despite the clear advantages of the dual-guide CRISPR-Cas13 platform in terms of improved specificity and mismatch discrimination, several limitations remain. First, the detection limit under the current experimental conditions was approximately 0.1 ng of genomic DNA, below which no signal was observed. This sensitivity, while sufficient for many diagnostic applications, may require further enhancement for samples with extremely low RNA abundance, such as liquid biopsies or early-stage infections. Second, although the system showed robust discrimination of single-nucleotide variants and successful SARS-CoV-2 detection, it has not yet been optimized for multiplexing, like conventional CRISPR platform [42]. Third, we still need to optimize the protocol to enable amplification-free detection, eliminating the reliance on PCR. This would not only streamline the workflow but also reduce potential bias and increase applicability in point-of-care settings.

In conclusion, this dual-guide CRISPR-Cas13 platform provides enhanced specificity and stable signal amplification for RNA detection. Its ability to discriminate single-nucleotide differences and reduce background activity holds promise for next-generation molecular diagnostics, with potential applications in infectious disease monitoring and precision oncology. In addition, these features could be further exploited for multiplexed diagnostic platforms or future applications in RNA editing and regulation.

## DATA AVAILABILITY

The data underlying this article will be shared on reasonable request to the corresponding author.

## SUPPLEMENTARY DATA

Supplementary Data are available at NAR online.

## AUTHOR CONTRIBUTIONS

A.A-G, J.J.D-M., F.M., I.M-J.: conceptualization and methodology. A.A-G, I.M-J, P.P-S: investigation and validation. A.A-G, I.M-J, J.J.D-M., F.M: formal analysis. A.A-G, I.M-J, I.R-H, F.J.M-E, P.P-S, S.R-P, R.T, R.S-M: writing-review & editing. R.M.S-M, J.J. D-M and F.M: Funding acquisition, project administration, resources and supervision. A.A-G, I.M-J., R.M.S-M, J.J.D-M., F.M: visualization, writing-original draft and review & editing. I. M-J. is a PhD student in Biomedicine at the University of Granada, and this work is part of his Doctoral Thesis. All authors have read and agreed to the published version of the manuscript.

## ACKNOWLEDGEMENTS

We thank Lorgen S.L. for their collaboration and support for the validation of our SARS-CoV-2 detection assays. We also express our gratitude to the Biobank of the Public Health System of Andalusia for the sample provision.

## FUNDING

This work was supported by the Spanish Ministry of Science and Innovation (MCIN)/AEI/10.13039/501100011033 and the European Union Next Generation EU/PRTR (Grant PID2022-141065OB-I00) as well as, by FEDER/Junta de Andalucía-Consejería de Economía y Conocimiento/Project CV20-77741. Additional support was provided by the Instituto de Salud Carlos III (ISCIII) through research projects PI21/00298 and PI24/00888, a TerAv (RD21/0017/0004) and TerAv+ (RD24/0014/0005]. By the Consejería de Salud y Familias (Junta de Andalucía) (PI-0236-2024). By the European Cooperation in Science and Technology (COST) [GeneHumdi-CA21113]. IMJ was supported by a predoctoral fellowship from the Spanish Ministry of Science, Innovation and Universities (FPU22/03455).

## CONFLICT OF INTEREST

Conflict of interest: F.M., A.A.G., J.J.D.M., and R.M.S.M are inventors of the patent Number: EP4414452A1, “DUAL-GUIDE RNA COMPOSITION FOR EXECUTING A SINGLE-GUIDE RNA CRISPR-ASSOCIATED SYSTEM”.

## Notes

### Author Declarations

The Research Ethics Committee of the Portal de Etica de la Investigacion Biomedica de Andalucia (PEIBA), Junta de Andalucia gave ethical approval for this work (Acta 6/2020, 29 June 2020). All saliva samples were provided by the Biobank of the Public Health System of Andalusia (reference S2000262), and informed consent was obtained from all participants.

