## Supplementary data for "A Novel Dual-Guide CRISPR-Cas13 Strategy Improves Specificity for Single-Nucleotide Variant Detection"

SUPLEMENTARY DATA FOR

### SUPPLEMENTARY MATERIAL AND METHODS

#### RT-RPA

RPA amplification had to be done using the TwistAmp® Basic kit (TwistDX Limited, Cambridge, UK) together with reverse transcription (ProtoScript® II Reverse Transcriptase, New England BioLabs, Ipswich, MA, USA) on the RNA samples using the system primers listed at **Supplementary Table S1**, following manufacturer's instructions.

**Supplementary Table S1. Oligonucleotide primers used in this study.**

| Name | Sequence (5′ - 3′) | Application |
|---|---|---|
| 2019-nCoV_N1-Fw | GACCCCAAAATCAGCGAAAT | qPCR |
| 2019-nCoV_N1-Rev | TCTGGTTACTGCCAGTTGAATCTG | |
| hGAPDH fw | TCAAGGCTGAGAACGGGAAG | qPCR |
| hGAPDH rev | CGCCCCACTTGATTTTGGAG | |
| Cov2_S1 Fw | GCCACTAGTCTCTAGTCAGTGTG | PCR and sequencing |
| Cov2_S1 Rev | TCAGGGTAATAAACACCACG | |
| S RPA Cov Fw | GAAATTAATACGACTCACTATAGGGGCAGCTTATTATGTGGGTTATTCTTCAAC | RPA |
| S RPA Cov Rev | GTACACTTTGTTTCTGAGAGAGGGTC | |
| KRAS Fw 1 | GATCCTTTGAGAGCCTTTAGC | PCR and sequencing |
| KRAS Rev 1 | GACCCTGACATACTCCCAAG | |
| sCoV Fw | GAAATTAATACGACTCACTATAGGGAGGTTTCAAACTTTACTTGCTTTACATAGA | PCR Sherlock |
| sCoV Rev | ACTTTGTTTCTGAGAGAGGGTCAAGTG | |
| KRAS T7 non-specific Fw | GAAATTAATACGACTCACTATAGGGGCCTGCTGAAAATGACTGAATATAA | PCR Sherlock |
| KRAS non-specific Rev | GTTGGATCATATTCGTCCACAAAA | |

**Supplementary Table S2. CRISPR-Cas13 guide RNAs used in this study.** The table lists all single and dual guide RNAs designed for the detection of SARS-CoV-2 and KRAS mutations.

| Name | Sequence (5′ - 3′) |
|---|---|
| sgRNA COVID | GAUUUAGACUACCCCAAAAACGAAGGGGACUAAAACUGUAAUGGUUCCAUUUUCAUUAUAUUUU |
| sgRNA COVID OFF5 | GAUUUAGACUACCCCAAAAACGAAGGGGACUAAAACUGUAUUGGUUCCAUUUUCAUUAUAUUUU |
| sgRNA COVID OFF3&5 | GAUUUAGACUACCCCAAAAACGAAGGGGACUAAAACUGAAUUGGUUCCAUUUUCAUUAUAUUUU |
| dtracrRNA dual 7 mer | GAUUUAGACUACCCCAAAAACGAAGGGGACUAAAACGACAGAUA |
| dtracrRNA dual 5 mer | GAUUUAGACUACCCCAAAAACGAAGGGGACUAAAACGAGCGC |
| dcRNA dual 7 mer COVID | UGUAAUGGUUCCAUUUUCAUUAUAUUUUUAUCUGU |
| dcRNA dual 5 mer COVID | UGUAAUGGUUCCAUUUUCAUUAUAUUUUGCGCU |
| dcRNA dual 5 mer COVID OFF5 | UGUAAUGGUUCCAUUUUCAUUAUUUUUUGCGCU |
| dcRNA dual 5 mer COVID OFF3&5 | UGUAAUGGUUCCAUUUUCAUUAUUUAUUGCGCU |
| sgrRNA KRAS WT | GAUUUAGACUACCCCAAAAACGAAGGGGACUAAAACCCUACGCCACCAGCUCCAACUACCACAA |
| sgrRNA KRAS G12D 19 | GAUUUAGACUACCCCAAAAACGAAGGGGACUAAAACGCACUCUUGCCUACGCCAUCAGCUCCAA |
| sgrRNA KRAS G12C 19 | GAUUUAGACUACCCCAAAAACGAAGGGGACUAAAACGCACUCUUGCCUACGCCACAAGCUCCAA |
| dcrRNA dual 5mer KRAS WT G12D 10 | GCACUCUUGCCUACGCCACCAGCUCCAAGCGCU |
| dcrRNA dual 5mer KRAS G12D 10 | GCACUCUUGCCUACGCCAUCAGCUCCAAGCGCU |
| dcrRNA dual 5mer KRAS G12C 10 | GCACUCUUGCCUACGCCACAAGCUCCAAGCGCU |

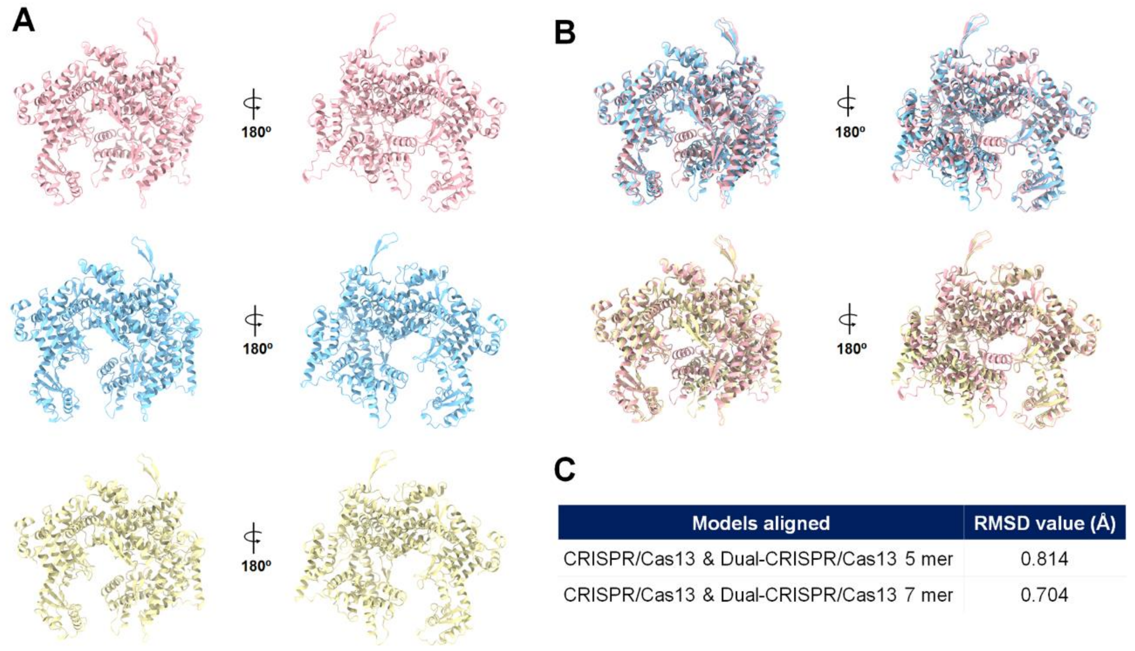


| Models aligned | RMSD value (Å) |
|---|---|
| CRISPR/Cas13 & Dual-CRISPR/Cas13 5 mer | 0.814 |
| CRISPR/Cas13 & Dual-CRISPR/Cas13 7 mer | 0.704 |

**Supplementary Figure S1. Structural comparison of Cas13a conformations in conventional and Dual-CRISPR systems using Alphafold 3.** A) Predicted structural models of LwaCas13a using AlphaFold 3. Models correspond to CRISPR (pink), Dual-CRISPR 5-mer (blue), and Dual-CRISPR 7-mer (yellow) configurations. For better clarity, the guide and target RNAs have been omitted. B) Structural superposition of each Dual-CRISPR model (blue and yellow) with the canonical CRISPR model (pink) reveals high structural similarity. C) Root-mean-square deviation (RMSD) values obtained from pairwise alignments of the Dual-CRISPR models with the conventional CRISPR model. RMSD values below 1 Å indicate high structural overlap and conformational similarity among the models.

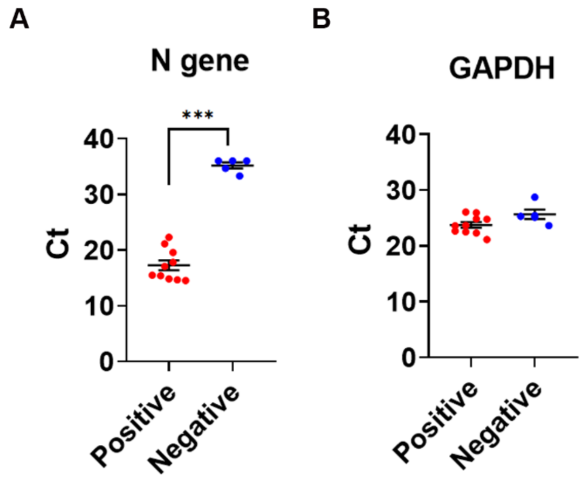


**Supplementary Figure S2. RT-qPCR analysis of SARS-CoV-2 N gene expression of different human samples.** Ct values for the viral N gene (A) and the endogenous control GAPDH (B) are shown for SARS-CoV-2-positive (patient's samples) and negative samples (healthy controls). Positive samples exhibited low Ct values for the N gene, indicating the presence of viral RNA, whereas healthy controls showed high or undetectable Ct values. GAPDH served as an internal control for RNA integrity and was consistently detected in all samples, with minor expected variation. Statistical analysis unpaired t-test, two-tails (* p < 0.05, ** p < 0.01, *** p < 0.001, **** p < 0.001). Values represent the mean ± SEM of at least 3 separate experiments.

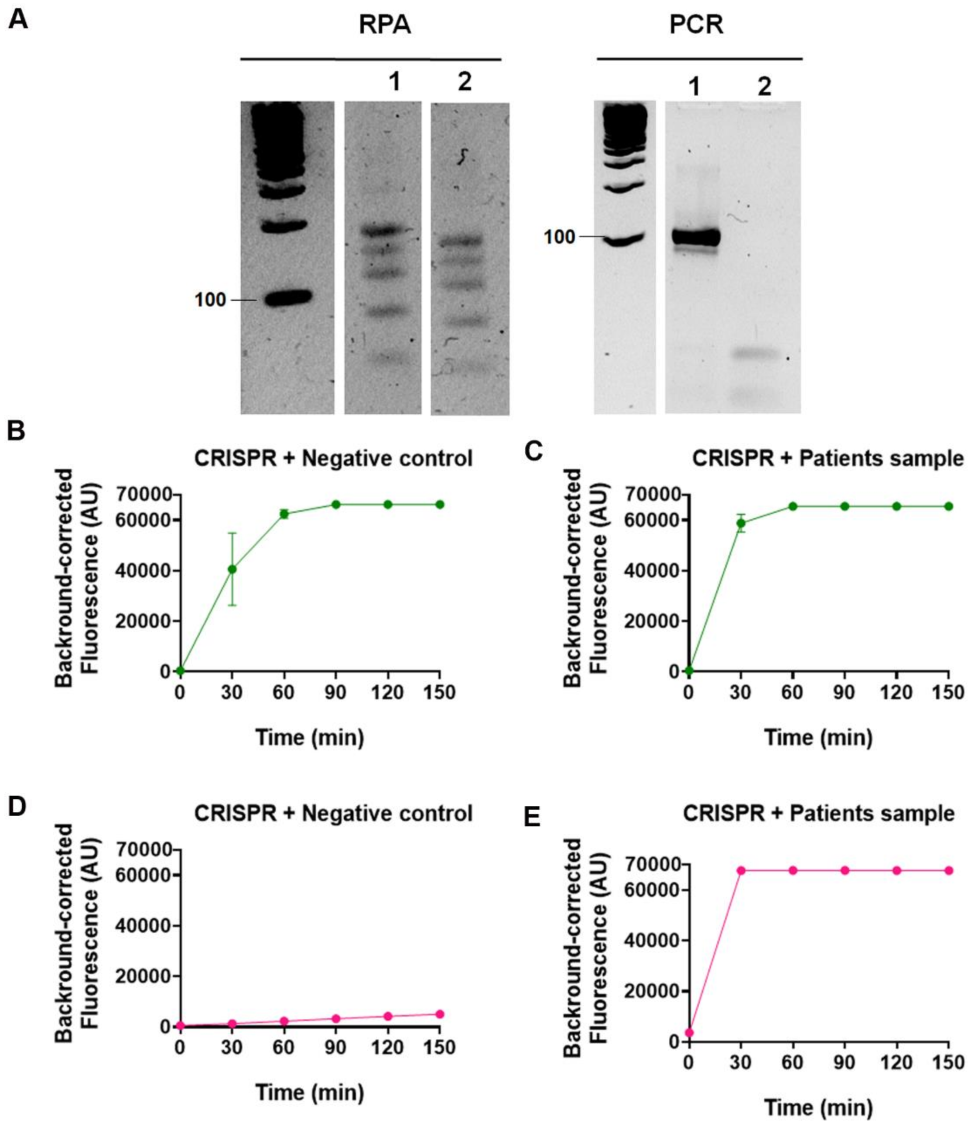


**Supplementary Figure S3. RPA generates false-positive signals due to non-specific amplification and cross-contamination.** A) Agarose gel electrophoresis of samples amplified by RPA or PCR. Sample 1 corresponds to a positive SARS-CoV-2 cDNA sample amplified with specific primers for RPA or PCR. Sample 2 corresponds to the negative control. A clear band of ~100 bp is expected for the positive sample, while the negative control should show no amplification. B–C) Corrected fluorescence of the CRISPR-Cas13 detection complex incubated with the fluorescent reporter for 150 min using RPA-amplified negative control (B) or positive patient samples (C). D–E) Corrected fluorescence of the CRISPR-Cas13 detection complex incubated with the fluorescent reporter for 150 min using PCR-amplified negative control (D) or positive patient samples (E).

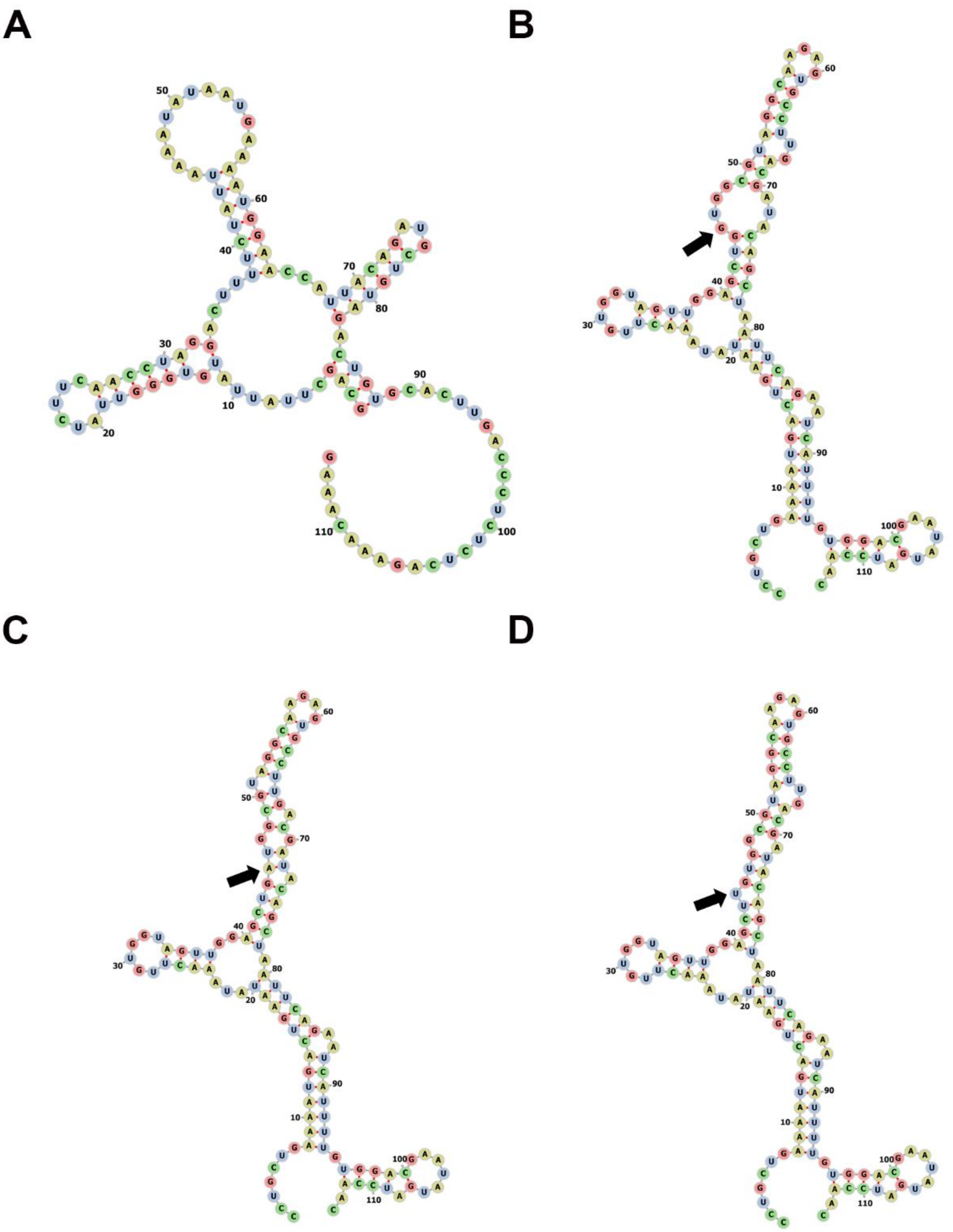


**Supplementary Figure S4. Predicted secondary structures of target RNAs used in CRISPR-Cas13 assays.** A) T7 in vitro transcribed RNA fragment from SARS-CoV-2, predicted to form three stem-loop domains with accessible single-stranded regions. B–D) T7 in vitro transcribed RNA fragments from the human KRAS gene: wild-type (B), G12D mutant (C), and G12C mutant (D). All three display elongated structures with multiple hairpins and internal loops. The mutated nucleotide is indicated with a black arrow. Structures were predicted using FoRNA software under default parameters.

**A**

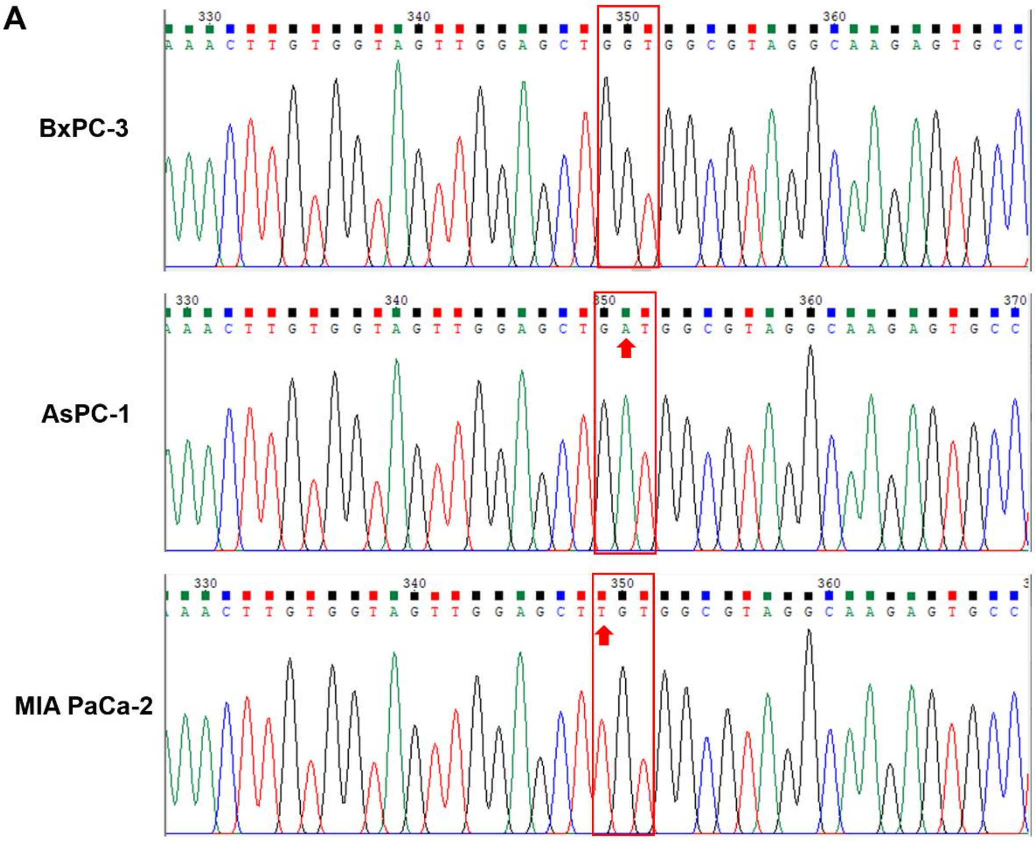


**B**

| Cell line | KRAS | Mutation |
|---|---|---|
| BxPC-3 | WT | None |
| AsPC-1 | G12D (homozygous) | GGT→GAT |
| MIA PaCa-2 | G12C (homozygous) | GGT→TGT |

**Supplementary Figure S5. Validation of KRAS gene mutations by sequencing in pancreatic cancer cell lines.** Codon 12 point mutations were identified in KRAS G12C (GGT→TGT) and G12D (GGT→GAT) variants, compared to the wild-type (WT) sequence. The pancreatic cancer cell lines BxPC-3, AsPC-1, and MIA PaCa-2 were analysed. All sequences were confirmed by Sanger sequencing.

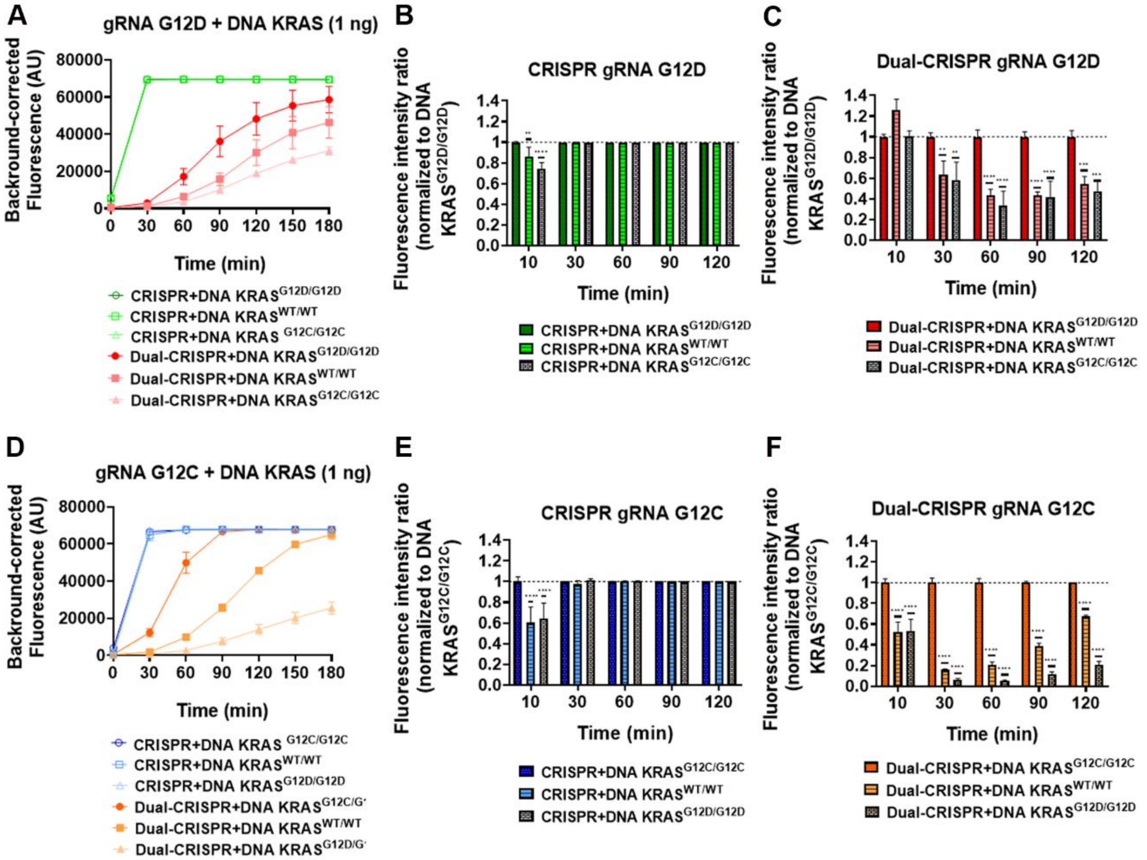


**Supplementary Figure S6. Sensitivity analysis of CRISPR and Dual-CRISPR/Cas13 systems for KRAS G12D and G12C detection using 1 ng of input DNA.** A) Corrected fluorescence of RNP-G12D guides (CRISPR and Dual-CRISPR) incubated with 1 ng of DNA from pancreatic cell lines harboring KRAS G12D, WT, or G12C alleles after 180 minutes. B) Fluorescence intensity ratio normalized to KRAS G12D DNA using RNP-G12D guides (CRISPR) at 30, 60, 90, and 120 minutes. C) Fluorescence intensity ratio normalized to KRAS G12D DNA using RNP-G12D guides (Dual-CRISPR) at 30, 60, 90, and 120 minutes. D) Corrected fluorescence of RNP-G12C guides (CRISPR and Dual-CRISPR) incubated with 1 ng of DNA from cell lines carrying KRAS G12C, WT, or G12D mutations after 180 minutes. E) Fluorescence intensity ratio normalized to KRAS G12C DNA using RNP-G12C guides (CRISPR) at 30, 60, 90, and 120 minutes. F) Fluorescence intensity ratio normalized to KRAS G12C DNA using RNP-G12C guides (Dual-CRISPR) at 30, 60, 90, and 120 minutes. Statistical analysis: Two-way ANOVA with Dunnett's multiple comparisons test was applied to compare specific targets vs. non-specific sequences at each time point (*p < 0.05, **p < 0.01, ***p < 0.001, ****p < 0.0001). Data represent mean ± SEM from three independent experiments.

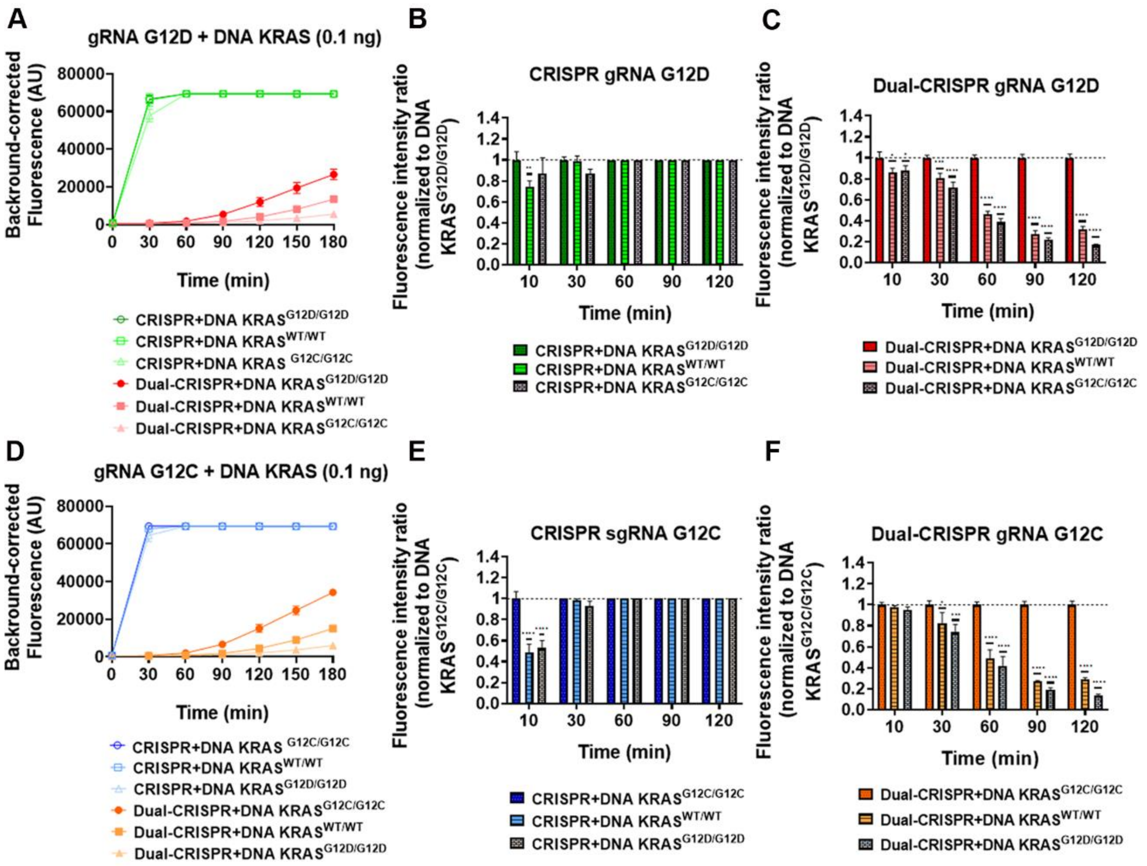


**Supplementary Figure S7. Ultra-sensitive detection of KRAS mutations using CRISPR and Dual-CRISPR/Cas13 systems with 0.1 ng of input DNA.** A) Corrected fluorescence of RNP-G12D guides (CRISPR and Dual-CRISPR) incubated with 0.1 ng of DNA from pancreatic cell lines harboring KRAS G12D, WT, or G12C alleles after 180 minutes. B) Fluorescence intensity ratio normalized to KRAS G12D DNA using RNP-G12D guides (CRISPR) at 30, 60, 90, and 120 minutes. C) Fluorescence intensity ratio normalized to KRAS G12D DNA using RNP-G12D guides (Dual-CRISPR) at 30, 60, 90, and 120 minutes. D) Corrected fluorescence of RNP-G12C guides (CRISPR and Dual-CRISPR) incubated with 0.1 ng of DNA from cell lines carrying KRAS G12C, WT, or G12D mutations after 180 minutes. E) Fluorescence intensity ratio normalized to KRAS G12C DNA using RNP-G12C guides (CRISPR) at 30, 60, 90, and 120 minutes. F) Fluorescence intensity ratio normalized to KRAS G12C DNA using RNP-G12C guides (Dual-CRISPR) at 30, 60, 90, and 120 minutes. Statistical analysis: Two-way ANOVA with Dunnett's multiple comparisons test was applied to compare specific targets vs. non-specific sequences at each time point (*$p < 0.05$, **$p < 0.01$, ***$p < 0.001$, ****$p < 0.0001$). Data represent mean ± SEM from three independent experiments.

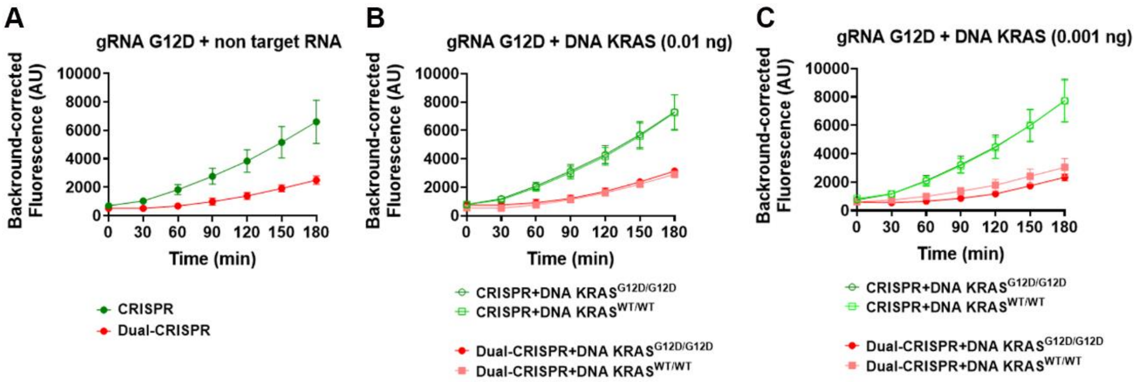


**Supplementary Figure S8. Evaluation of the detection limit of the Dual-CRISPR-Cas13 system using decreasing amounts of target input.** The system was tested with no target RNA (A) and 0.01 ng (B), and 0.001 ng (C) of target nucleic acid. There was no differences at fluorescence level from the non-target controls (A) with the complex corrected fluorescence of RNP-G12D guides (CRISPR and Dual-CRISPR) incubated with 0.01 ng (B) and 0.001 ng (C) of DNA from pancreatic cell lines harboring KRAS G12D and WT alleles after 180 minutes.
